# Healthcare-Associated COVID-19 in Ontario, Canada: Relative Mortality and Contribution to Community Epidemic Growth

**DOI:** 10.1101/2025.07.28.25332157

**Authors:** Natalie J. Wilson, Alicia A. Grima, Clara Eunyoung Lee, David N. Fisman

**Affiliations:** Dalla Lana School of Public Health, University of Toronto, Toronto ON, Canada

## Abstract

**Background:** The COVID-19 pandemic placed immense strain on Canada’s healthcare system and disproportionately affected individuals with poorer baseline health. Healthcare-associated infections (HAIs) increase risk for both patients and healthcare workers and are often more severe due to advanced age and comorbidities. While efforts have aimed to reduce in-hospital transmission, the individual- and community-level consequences of HAIs require further study. We aimed to assess whether healthcare-associated COVID-19 cases had higher odds of death compared to hospitalized community-acquired cases, and to evaluate the directionality of transmission between hospitals and the community.

**Methods:** We analyzed COVID-19 surveillance data from Ontario’s Case Contact and Management System and the COVaxON vaccine registry (March 17, 2020, to September 4, 2022). Latent class analysis was used to classify hospitalized cases by likelihood of healthcare-associated infection. Mortality odds by category were estimated using binomial logistic regression. Directionality between hospital outbreaks and community cases was assessed using a modified Granger causality approach.

**Findings:** Compared to patients with low likelihood of healthcare-associated infection, those moderately likely to have acquired COVID-19 in hospital had elevated odds of death (OR: 1.26, 95% CI: 1.14–1.40); no significant increase was seen in the high-likelihood group (OR: 1.05, 95% CI: 0.96–1.15). Community cases did not predict hospital outbreaks (p=0.5749), but hospital outbreaks predicted community case growth (p<0.0001).

**Interpretation:** Hospital-acquired COVID-19 is associated with excess mortality and may drive community transmission. Preventing in-hospital transmission is critical to protecting patients and controlling broader epidemic spread.

**Funding:** Supported by a Canadian Institutes for Health Research project grant, #518192.

## Introduction

The first case of severe acute respiratory syndrome coronavirus 2 (SARS-CoV-2) in Ontario, Canada was detected in late January of 2020. ^1^ In the following months and years, coronavirus disease 2019 (COVID-19) has placed immense strain on the Canadian healthcare system and become one of the leading causes of death nationwide. ^2,3^

Despite infection control efforts, healthcare settings present a pathway for SARS-CoV-2 transmission. ^4,5^ During early stages of the pandemic, hospitals in Canada and globally faced challenges in controlling SARS-CoV-2 transmission due to high patient volumes, limited testing capacity, and high viral transmissibility. Clinical characteristics of COVID-19 vary across individuals, however, pre-existing comorbidities are associated with increased mortality and disease severity, resulting in a disproportionate impact on elderly patients and those with poorer baseline levels of health. ^6^ Nosocomial viral transmission impacts patients who often already have risk factors associated with severe outcomes, puts healthcare workers at risk for infection, and creates excess pressure on the healthcare system. Previous studies on relative mortality in patients with healthcare-acquired compared to non-healthcare acquired COVID-19 have had mixed findings. ^7–9^ However, many existing studies only examined case data from before SARS-CoV-2 vaccines were available and analyzed hospital-level rather than population-level data. Nosocomial transmission of SARS-CoV-2 can lead to outbreaks in hospital settings, and patients with hospital-acquired SARS-CoV-2 have been found to be the main source of hospital transmission to other patients. ^10^

Not only does nosocomial transmission negatively impact affected patients, the dynamics of transmission between healthcare settings and the broader community are not yet fully understood. Healthcare facilities are strongly interconnected with the communities in which they exist, through staff, family members, and patients entering and exiting the facility, so it is important to consider the mutual influence these environments have on each other. Hospital transmission has been previously shown to play a significant role in the spread of Middle East respiratory syndrome coronavirus (MERS-CoV) and severe acute respiratory syndrome coronavirus 1 (SARS-CoV-1).^11,12^ When punctuated lockdowns are implemented as a pandemic control measure, hospital transmission may have a significant influence on community spread despite making up a small proportion of overall cases. ^10^ Understanding the interconnection between hospital and community transmission has implications for infection prevention and control practices as new viral variants continue to emerge, and these findings can be applied to novel public health threats in the future.

In this study, we investigated whether healthcare-associated cases had higher odds of death from COVID-19 compared to hospitalized community-acquired cases, and explored directionality of COVID-19 case counts between the community and hospital outbreaks.

## Methods

### Data collection

We obtained COVID-19 surveillance data in Ontario, Canada including case counts, outbreak information, hospitalizations, Public Health Unit (PHU) information, and vaccination coverage. Data was obtained from the Ontario Case Contact and Management (CCM) System and the COVaxON vaccine information system. CCM data includes laboratory-confirmed COVID-19 cases derived from case records from Ontario’s PHUs. ^13^ CoVaxON vaccination data was linked to CCM case data through a unique identifier used in both databases to associate cases with their individual vaccination status. ^14^ Population denominators were obtained from Statistics Canada. ^15^ COVID-19 test and hospitalization data was available from March 17, 2020, to September 4, 2022.

### Healthcare-associated case analysis

COVID-19 cases identified as being hospitalized were categorized based on the time interval between their hospital admission and their positive COVID-19 test date. Patients with a PCR-confirmed COVID-19 infection 2 days or less after admission were considered community-acquired; 3–7 days after admission were considered indeterminate healthcare-associated; 8–14 days after admission were considered probable healthcare-associated; and 15 days or more after admission were considered definite healthcare-associated as defined by Cooper *et al*. ^10^

### Latent class analysis

There are several possible approaches to assigning a linkage between healthcare exposure and acquisition of SARS-CoV-2. CCM data contain a “nosocomial” flag; cases might also be identified as likely healthcare-acquired if they are hospitalized cases linked to an assigned SARS-CoV-2 outbreak number; lastly, cases might be identified as healthcare acquired if first testing date for SARS-CoV-2 occurs at some lag after hospitalization. To incorporate each of the various definitions of healthcare-acquired SARS-CoV-2 infections, we used latent class analysis (LCA), which uses a probabilistic approach to identify unobserved subgroups (latent classes) based on patterns in indicator variables. ^16^ Our LCA was based on five binary indicator variables: nosocomial classification in CCM, hospital-outbreak associated cases, positive SARS-CoV-2 test 3+ days after hospital admission, positive test 8+ days after admission, and positive test 15+ days after admission. The LCA model assigns each case to its most likely class based on the covariance of the five indicator variables to form an optimal number of classes. We tested two, three, and four class approaches, but a three-class solution was selected due to having the lowest Bayesian information criterion. We then classified the three classes as unlikely healthcare-acquired, moderately likely healthcare-acquired, and likely healthcare-acquired. Odds of death in hospitalized patients based on likelihood of healthcare-associated infection from the LCA were analyzed using binomial logistic regression. Covariates included age, presence of comorbidities, immunocompromised host (ICH), long-term care (LTC) home residence, vaccination status, and pandemic wave. Patients were considered to be vaccinated against COVID-19 if they had received at least two doses of an approved COVID-19 vaccine at least 14 days prior to their positive test. Pandemic waves were defined based on the dominant SARS-CoV-2 variant at the time in Ontario using the approach previously described by Mitchell *et al*. ^17^ Briefly, cases in wave one occurred before August 31, 2020 (wild-type variant), wave two cases occurred from September 1, 2020 to February 28, 2021 (wild-type variant), wave three occurred from March 1 to June 30, 2021 (Alpha, Beta, and/or Gamma variants), wave four occurred from July 1 to December 25, 2021 (Delta variant), wave five occurred from December 26, 2021 to March 19, 2022 (Omicron variant), and wave six included cases occurring after March 20, 2022 (Omicron variant). Cases that were never hospitalized were not included in the analysis. The LCA was also used to estimate sensitivity and specificity of each nosocomial infection indicator variable, with the moderately likely and likely healthcare-acquired groups acting as “cases”, and the unlikely healthcare-acquired group acting as “non-cases”.

### Hospital-associated outbreak and community case forecasting

To evaluate the directionality between hospital-associated COVID-19 outbreaks and community cases, we used a modified Granger causality approach. Typically, Granger causality uses vector autoregression models for continuous data, but to apply this concept to count data we used a Poisson regression model with lagged predictor variables. We created lagged variables for hospital outbreak case counts and test-adjusted community case counts 2 weeks preceding the reported case and outbreak counts for each of Ontario’s 34 PHUs. Test-adjusted case counts are estimated via a regression-based approach that accounts for the profound ascertainment bias in Ontario case data, with individuals at elevated risk of hospitalization and mortality based on age are tested at far higher rates (and consequently over-represented in case data) than individuals at lower risk. ^13^ This method has been validated as generating case rates that predict mortality more accurately than crude reported case rates. ^18^

Hospital outbreak-associated cases included all cases of COVID-19 flagged as being associated with a hospital outbreak, regardless of individual hospitalization status. If inclusion of the lagged variable in the model significantly improves fit in the dependent variable, it suggests that the lagged variable forecasts the outcome. We conducted fixed effects Poisson regression models using hospital outbreak case counts as the dependent variable and 2-week lagged test-adjusted community case counts as the independent variable, and vice versa. Fixed effects were included for public health unit and COVID-19 vaccination coverage was included as a covariate.

Vaccination coverage was calculated as cumulative COVID-19 vaccine doses administered per 100,000 residents in each PHU.

For hospitalized cases:

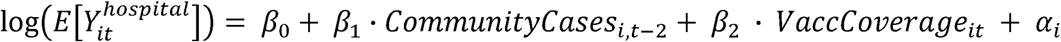

For community cases:

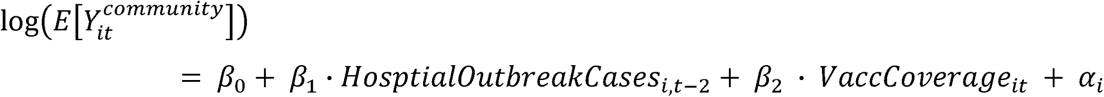

Subscripts (*i,t*) represent *i^th^* PHU. The term log(E[Y]) is the predicted case rate. The term α_i_ represents PHU fixed effects.

We used a Wald test to test whether inclusion of the lagged predictor variable significantly improved the model fit and may forecast future community cases or hospital outbreak-related cases.

Analyses were performed using Stata version 15.1 and R Studio 2024.12.1+563. This study was approved by the University of Toronto Health Sciences Research Ethics Board under Protocols #46160. The analysis involved secondary use of data derived from provincial surveillance and health system databases. Individual-level records used for latent class analysis, logistic regression and data aggregation contained no personal health identifiers, and no individual consent was required. This study adhered to the principles of the Declaration of Helsinki. The funders had no role in study design, data collection, data analysis, data interpretation, or writing of the report. The corresponding author had full access to all the data and had final responsibility for the decision to submit for publication.

## Results

### Hospital outbreak characteristics

From the onset of COVID-19 outbreak reporting in Ontario until the end of data availability, a total of 21,862 COVID-19 outbreaks in all settings were identified. There were 1,838 hospital outbreaks which were associated with 17,785 cases. The number of hospital outbreaks per 100,000 residents in each PHU in Ontario is displayed in **Figure 1**.

**Figure 1:**
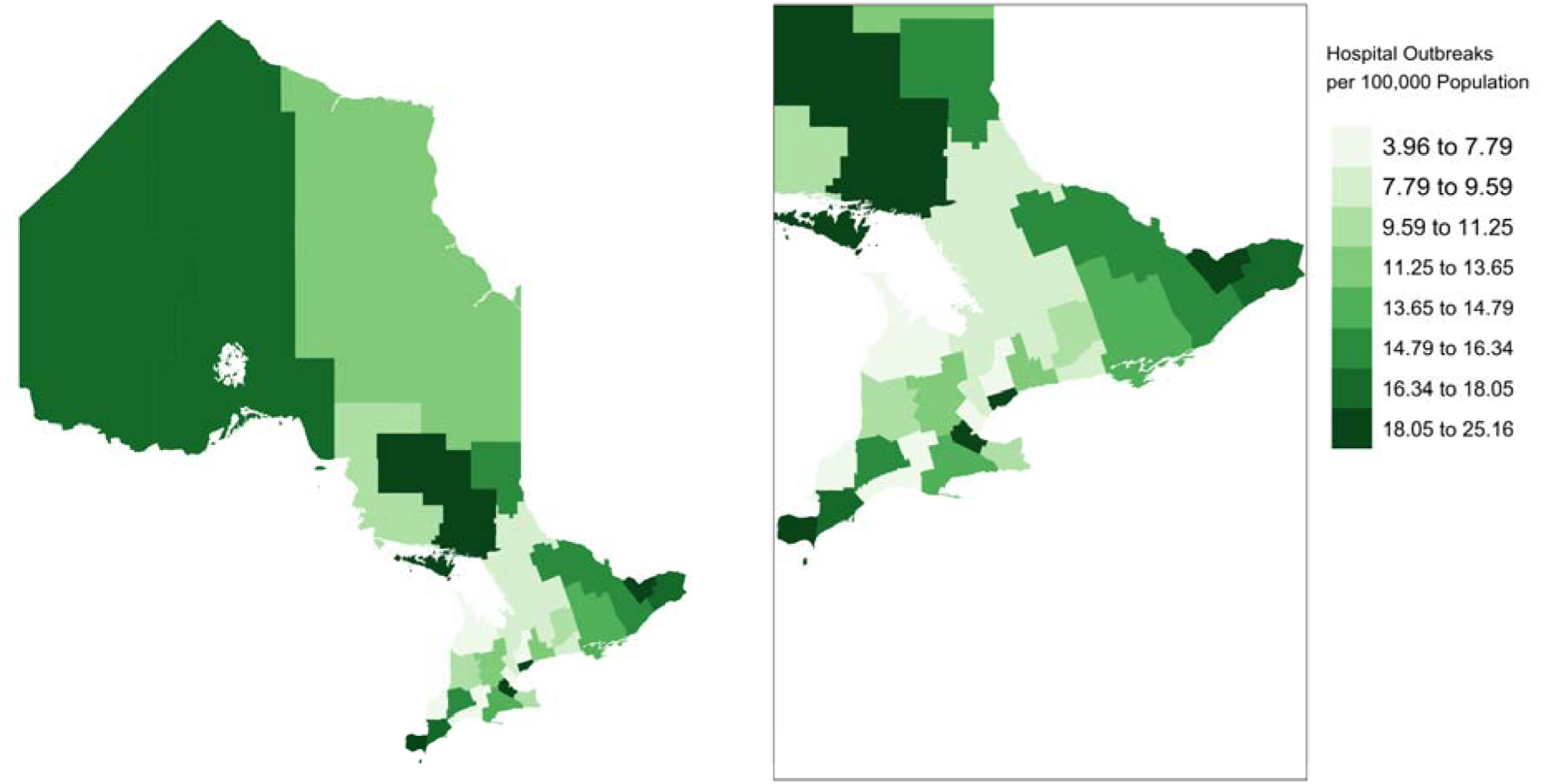
Map of Ontario showing cumulative hospital-associated SARS-CoV-2 outbreak count per 100,000 residents by Public Health Unit from March 17, 2020, through September 4, 2022.

### Healthcare-associated case analysis

A total of 53,665 cases of COVID-19 were hospitalized between March 17, 2020, and September 4, 2022. 53,191 cases had complete data and were included in our analysis. Of hospitalized cases, 13·89% were linked to hospital outbreaks. Based on the time from hospital admission to positive COVID-19 test, 47,893 cases were classified as community-acquired, 1,613 cases were indeterminate healthcare-associated, 1,356 cases were probable healthcare-associated, and 2,329 cases were definite healthcare-associated (**Table 1**).

**Table 1:**
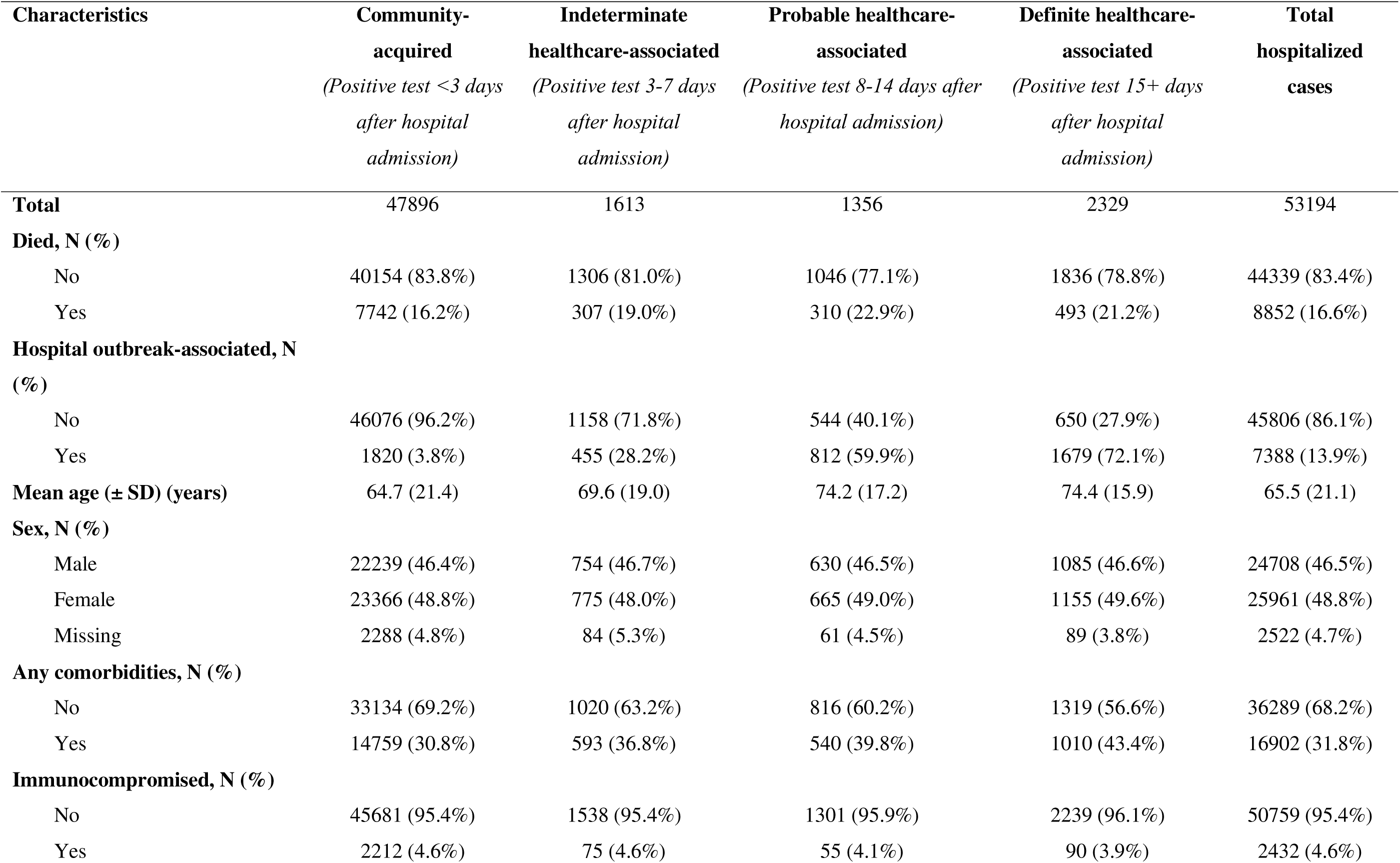
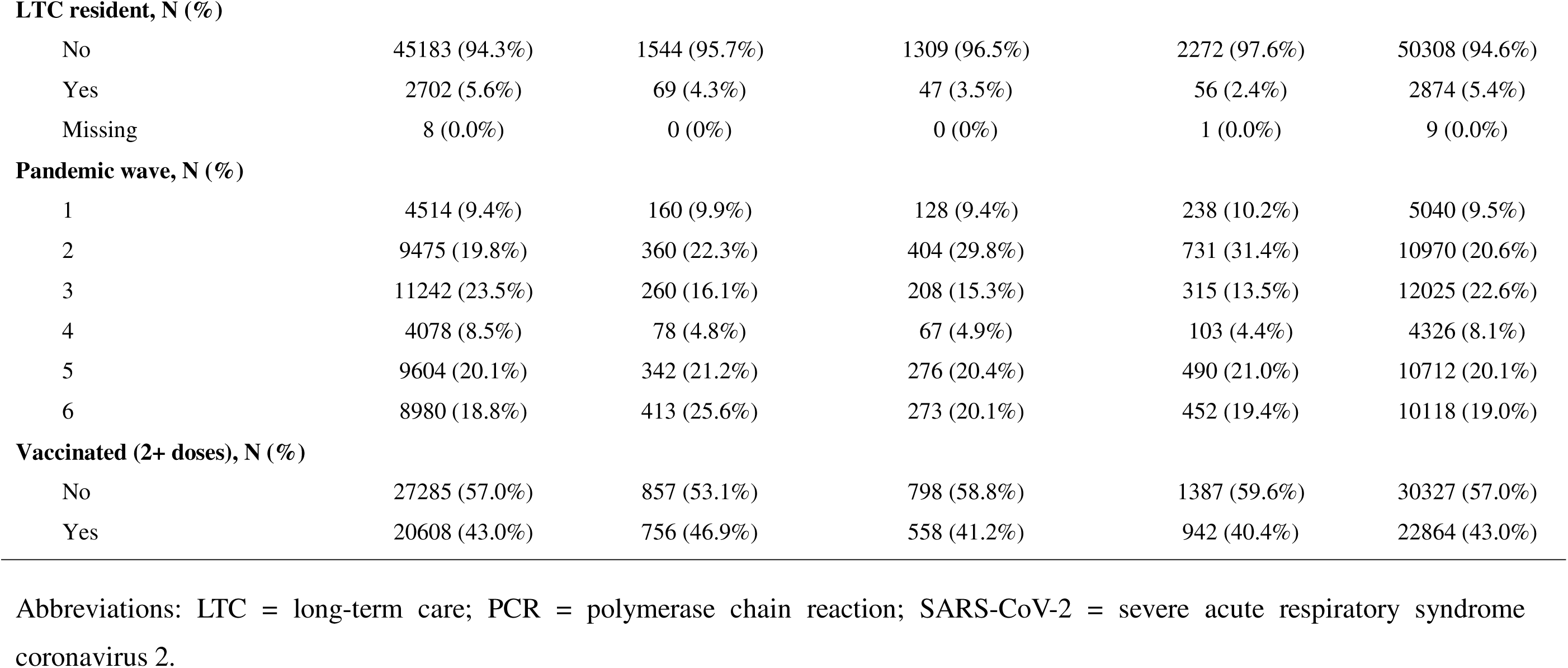
Characteristics of hospitalized COVID-19 cases based on time between hospital admission and positive SARS-CoV-2 PCR test.

| Characteristics | Community-acquired<br>*(Positive test <3 days*<br>*after hospital*<br>*admission)* | Indeterminate<br>healthcare-associated<br>*(Positive test 3-7 days*<br>*after hospital*<br>*admission)* | Probable healthcare-associated<br>*(Positive test 8-14 days after*<br>*hospital admission)* | Definite healthcare-associated<br>*(Positive test 15+ days*<br>*after hospital*<br>*admission)* | Total<br>hospitalized<br>cases |
| --- | --- | --- | --- | --- | --- |
| **Total** | 47896 | 1613 | 1356 | 2329 | 53194 |
| **Died, N (%)** |  |  |  |  |  |
| No | 40154 (83.8%) | 1306 (81.0%) | 1046 (77.1%) | 1836 (78.8%) | 44339 (83.4%) |
| Yes | 7742 (16.2%) | 307 (19.0%) | 310 (22.9%) | 493 (21.2%) | 8852 (16.6%) |
| **Hospital outbreak-associated, N (%)** |  |  |  |  |  |
| No | 46076 (96.2%) | 1158 (71.8%) | 544 (40.1%) | 650 (27.9%) | 45806 (86.1%) |
| Yes | 1820 (3.8%) | 455 (28.2%) | 812 (59.9%) | 1679 (72.1%) | 7388 (13.9%) |
| **Mean age (± SD) (years)** | 64.7 (21.4) | 69.6 (19.0) | 74.2 (17.2) | 74.4 (15.9) | 65.5 (21.1) |
| **Sex, N (%)** |  |  |  |  |  |
| Male | 22239 (46.4%) | 754 (46.7%) | 630 (46.5%) | 1085 (46.6%) | 24708 (46.5%) |
| Female | 23366 (48.8%) | 775 (48.0%) | 665 (49.0%) | 1155 (49.6%) | 25961 (48.8%) |
| Missing | 2288 (4.8%) | 84 (5.3%) | 61 (4.5%) | 89 (3.8%) | 2522 (4.7%) |
| **Any comorbidities, N (%)** |  |  |  |  |  |
| No | 33134 (69.2%) | 1020 (63.2%) | 816 (60.2%) | 1319 (56.6%) | 36289 (68.2%) |
| Yes | 14759 (30.8%) | 593 (36.8%) | 540 (39.8%) | 1010 (43.4%) | 16902 (31.8%) |
| **Immunocompromised, N (%)** |  |  |  |  |  |
| No | 45681 (95.4%) | 1538 (95.4%) | 1301 (95.9%) | 2239 (96.1%) | 50759 (95.4%) |
| Yes | 2212 (4.6%) | 75 (4.6%) | 55 (4.1%) | 90 (3.9%) | 2432 (4.6%) |
| **LTC resident, N (%)** |  |  |  |  |  |
| No | 45183 (94.3%) | 1544 (95.7%) | 1309 (96.5%) | 2272 (97.6%) | 50308 (94.6%) |
| Yes | 2702 (5.6%) | 69 (4.3%) | 47 (3.5%) | 56 (2.4%) | 2874 (5.4%) |
| Missing | 8 (0.0%) | 0 (0%) | 0 (0%) | 1 (0.0%) | 9 (0.0%) |
| **Pandemic wave, N (%)** |  |  |  |  |  |
| 1 | 4514 (9.4%) | 160 (9.9%) | 128 (9.4%) | 238 (10.2%) | 5040 (9.5%) |
| 2 | 9475 (19.8%) | 360 (22.3%) | 404 (29.8%) | 731 (31.4%) | 10970 (20.6%) |
| 3 | 11242 (23.5%) | 260 (16.1%) | 208 (15.3%) | 315 (13.5%) | 12025 (22.6%) |
| 4 | 4078 (8.5%) | 78 (4.8%) | 67 (4.9%) | 103 (4.4%) | 4326 (8.1%) |
| 5 | 9604 (20.1%) | 342 (21.2%) | 276 (20.4%) | 490 (21.0%) | 10712 (20.1%) |
| 6 | 8980 (18.8%) | 413 (25.6%) | 273 (20.1%) | 452 (19.4%) | 10118 (19.0%) |
| **Vaccinated (2+ doses), N (%)** |  |  |  |  |  |
| No | 27285 (57.0%) | 857 (53.1%) | 798 (58.8%) | 1387 (59.6%) | 30327 (57.0%) |
| Yes | 20608 (43.0%) | 756 (46.9%) | 558 (41.2%) | 942 (40.4%) | 22864 (43.0%) |

Abbreviations: LTC = long-term care; PCR = polymerase chain reaction; SARS-CoV-2 = severe acute respiratory syndrome coronavirus 2.

### Nosocomial infection indicators latent class analysis

Three latent classes were generated based on the five binary indicator variables: nosocomial classification in CCM, hospital-outbreak associated cases, positive SARS-CoV-2 test 3+ days after hospital admission, positive test 8+ days after admission, and positive test 15+ days after admission. These classes represent unlikely healthcare-acquired, moderately likely healthcare-acquired, and likely healthcare-acquired cases. Of 53,191 total hospitalized cases, 46,819 were classified as unlikely, 2,687 were classified as moderately likely, and 3,685 were classified as likely healthcare-acquired through the LCA. A breakdown of the intersection of indicator variables of hospitalized patients used in the LCA is presented in Figure 2. Compared to unlikely healthcare-associated cases, moderately likely, and likely healthcare-associated cases had 1·26 (95% CI: 1·14-1·40) and 1·05 (95% CI: 0·96-1·15) times the odds of death respectively after controlling for age, gender, presence of comorbidities, ICH, long-term care home residence, vaccination status, and pandemic wave. Estimated sensitivity and specificity of each indicator variable is presented in **Table 2**.

**Figure 2:**
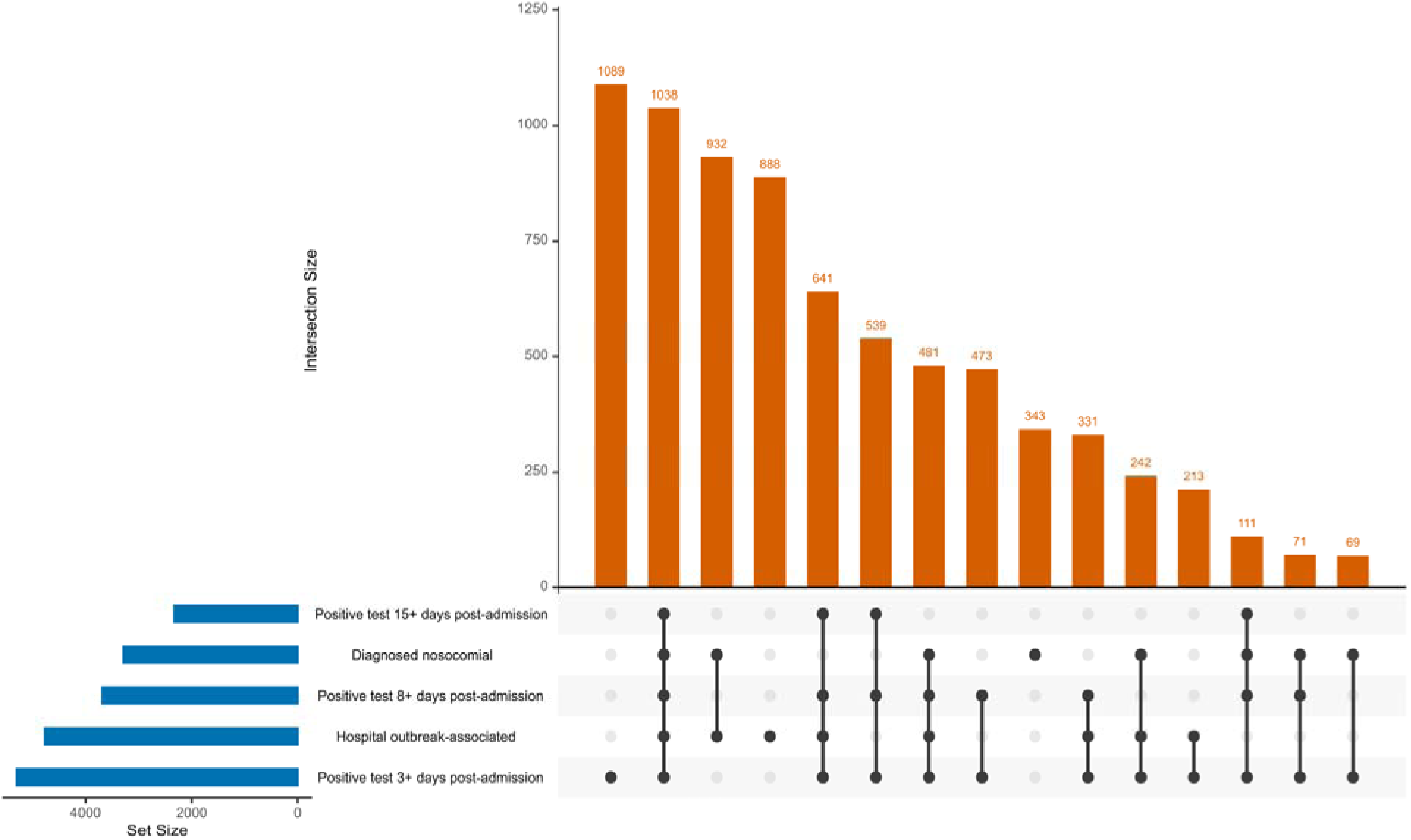
Intersection between five indicator variables used in latent class analysis of 53,191 total patients hospitalized with SARS-CoV-2.

**Table 2:** Sensitivity and specificity of nosocomial SARS-CoV-2 indicator variables as estimated through latent class analysis.

| Indicator variable | Sensitivity | Specificity |
| --- | --- | --- |
| Nosocomial classification in CCM | 0.705 | 1.000 |
| Hospital outbreak-associated | 0.486 | 1.000 |
| Positive test 3+ days after admission | 0.634 | 0.978 |
| Positive test 8+ days after admission | 0.545 | 1.000 |
| Positive test 15+ days after admission | 0.345 | 1.000 |

Abbreviations: CCM = Ontario COVID-19 case and contact management system; SARS-CoV-2 = severe acute respiratory syndrome coronavirus

### Hospital-associated outbreak and community case forecasting

Test-adjusted community cases with a 2-week lag did not improve forecasting of hospital outbreak cases (p=0·5749). However, hospital outbreak-associated cases with a 2-week lag did improve forecasting of test-adjusted community cases (p<0·0001).

## Discussion

In this population-based study of hospitalized COVID-19 cases in Ontario, we found that nosocomial SARS-CoV-2 infection is important for two distinct reasons, both of which challenge prevailing assumptions. First, patients moderately likely to have acquired COVID-19 in-hospital had significantly higher odds of in-hospital death than likely community-acquired cases among individuals sick enough to require hospitalization. This finding runs counter to the expectation that patients hospitalized for COVID-19 would represent those most at risk of mortality from the virus. Instead, our results suggest that acquiring COVID-19 during hospitalization confers even greater risk, presumably due to multiplicative risk in individuals who acquire this viral infection in combination with whatever health issue resulted in hospitalization. Patients classified as likely to have acquired SARS-CoV-2 did not have significantly elevated odds of in-hospital death compared to unlikely healthcare-associated cases, which is likely due to selection bias associated with healthier patients surviving long enough after hospital admission to contract and be diagnosed with COVID-19 before discharge or death. Second, we found that hospital outbreak-associated cases were predictive of subsequent increases in community case rates, whereas community case rates did not predict hospital outbreaks. This suggests that hospitals may not simply reflect community transmission trends but instead act as amplifiers of epidemic activity. Nosocomial COVID-19 is therefore not only dangerous for individual patients, but may also contribute disproportionately to wider community spread.

The reported relative severity of nosocomial COVID-19 has varied across studies. ^7–9^ The COPE-Nosocomial study, for example, reported lower adjusted mortality among hospital-onset cases using time-to-event analysis. However, that study applied a highly restrictive definition of hospital-acquired infection, requiring that diagnosis occur more than 14 days after admission. Our own data show that this definition yields perfect specificity but a sensitivity of only 34 percent, meaning it misses the majority of true nosocomial cases. In addition to low sensitivity, this approach introduces survivorship bias by conditioning case classification on remaining in hospital for at least 15 days without diagnosis or death. Patients who acquire COVID-19 during hospitalization but deteriorate rapidly and die before day 15 are systematically excluded from the nosocomial group, resulting in an artificially low-risk cohort and downwardly biased estimates of mortality.

Other frequently applied surveillance cutoffs, such as three- or eight-days post-admission, present different trade-offs: the three-day rule has moderate sensitivity, while the eight-day rule improves specificity at the cost of lower sensitivity. These findings underscore the limitations of single-indicator definitions and the risk of misclassifying cases in a way that would underestimate the relative severity of hospital-acquired infection by biasing effects toward the null.

In contrast, our study uses latent class modeling to probabilistically classify nosocomial infections based on multiple indicators. This approach avoids the sensitivity and specificity pitfalls of fixed cutoffs and captures a fuller spectrum of hospital-acquired infections. The resulting classification reveals a consistent gradient of elevated mortality risk corresponding to the likelihood of nosocomial acquisition, suggesting that hospital-acquired COVID-19 is not only more deadly but also more common than strict definitions imply. Our findings are consistent with observations by Melançon and Ponsford. ^7,8^ However, the Melançon analysis was limited to a single center and stratified results by age without extensive adjustment for other confounders, while the Ponsford study relied on hospital-level aggregate data and time-based definitions without individual-level multivariable control. By comparison, our study integrates individual-level data with robust adjustment and refined classification, providing stronger evidence that nosocomial infection contributes directly to poor outcomes, rather than simply reflecting underlying vulnerability.

Beyond elevated individual-level risk, our findings carry important implications for understanding epidemic dynamics. The observation that hospital outbreak-associated cases predict subsequent increases in community transmission challenges the prevailing assumption that hospitals are simply downstream recipients of infection. Instead, our results suggest that healthcare settings may function as amplifiers, transmitting infection outward through staff, discharged patients, or visitors. This directional pattern is consistent with modeling studies from England, ^4^ which showed that nosocomial cases contributed meaningfully to sustaining community transmission during periods when other transmission routes were constrained. The role of hospitals as amplifiers of infection transmission for other beta-coronaviruses, such as SARS-CoV-1 and MERS-CoV, is well established. ^11,12,19^ Recognizing hospitals as potential sources rather than just sinks of epidemic activity reframes infection control as not only an institutional safety measure, but also a critical component of broader public health strategy.

These findings point to the need for a fundamental rethinking of how in-hospital transmission is measured, interpreted, and prevented. Case definitions that emphasize specificity over sensitivity (e.g., 14-day cut-off) may simplify reporting but fail to reflect the true scope and consequences of nosocomial infection. When most hospital-acquired cases go undetected, both surveillance systems and institutional responses are weakened. Underestimating this burden not only distorts our understanding of disease dynamics but also reduces accountability for preventable harm within healthcare settings.

At the same time, infection prevention policies continue to lag behind the science of airborne transmission. ^20^ Many current protocols are still based on outdated droplet models and fail to provide meaningful protection against aerosolized pathogens. Effective prevention requires structural changes that include access to high-filtration respirators, improvements in ventilation, portable air cleaning, and routine monitoring of indoor air quality. ^21,22^ These measures are not aspirational. They are technically feasible, and they represent a modern standard of care.

Hospitals are not only places of care but also hubs of connectivity within healthcare systems and the wider community. As highly trafficked and densely networked institutions, they exert substantial gravitational pull in the spread of infection. ^23^ Patients, staff, and visitors move between hospitals and surrounding communities in ways that can accelerate regional transmission. This pattern, which we aim to explore further in future work, suggests that outbreaks within hospitals may serve as early indicators of community risk and as drivers of wider epidemic momentum. Ensuring clean, safe air and more responsive surveillance in these high-gravity settings is not just about protecting vulnerable individuals. It is essential for reducing epidemic potential at the population level.

This study has several limitations. First, as with all observational research, we cannot exclude the possibility of residual confounding, despite adjusting for a wide range of clinical and demographic covariates. Second, while our use of latent class modeling offers a more nuanced and probabilistic approach to nosocomial classification, it depends on the quality and completeness of input indicators, which may vary across institutions or over time. Third, our findings are based on acute care hospitals in Ontario and may not generalize to settings with different infection control practices, healthcare infrastructure, or patient characteristics. Fourth, although our ecological analysis suggests that hospital outbreaks precede increases in community transmission, we cannot definitively establish directionality. Even with individual-level linkage data, confirming the sequence of transmission would require viral genotyping, ^24^ which was not available. Despite these constraints, we believe that carefully constructed analyses using the best available data remain essential for guiding policy, particularly in rapidly evolving or resource-limited contexts.

Our findings challenge common assumptions about the relative risk and public health role of hospital-acquired SARS-CoV-2 infection. Consistent with prior work, we find that these infections are both more deadly for patients and more influential in shaping epidemic dynamics than current surveillance and prevention frameworks suggest. By using probabilistic classification and linking nosocomial case incidence to downstream community trends, we provide evidence that hospitals are not only vulnerable to SARS-CoV-2 transmission but can also act as accelerators of broader spread. Addressing this challenge requires both conceptual and practical shifts: moving beyond static case definitions and embracing airborne prevention as a core pillar of healthcare safety. Recognizing and responding to the full impact of hospital-acquired SARS-CoV-2 infection is essential for protecting individuals, strengthening health systems, and reducing population-level risk.

## Data Availability

Data Availability
The authors are bound by data-sharing agreements that preclude public release of individual-level surveillance data. Aggregated case data are available upon request to the corresponding author and may be shared in accordance with relevant institutional and data provider restrictions. Researchers may also contact the Ontario Ministry of Health to request access to primary datasets.

